# Life’s Essential 8 in Pregnancy and Time to Incident Cardiometabolic Disease Over 7 Years Follow-Up

**DOI:** 10.64898/2026.04.14.26350869

**Authors:** Ellen C. Francis, Shalin Patel, Anushka Pande, Alexa Freedman, Lauren Keenan-Devlin, Linda M. Ernst, Emily S. Barrett, Ann Borders, Gregory E. Miller, Shristi Rawal, Amy H. Crockett

## Abstract

**Importance:** Assessment of cardiovascular health (CVH) during may unmask latent metabolic vulnerability and indicate long-term disease risk. However, the prognostic value of the AHA’s Life’s Essential 8 (LE8) framework during pregnancy remains uncertain.

**Objective:** To evaluate CVH during using a modified Life’s Essential 8 (mLE8) score in association with time to incident cardiometabolic disease.

**Design:** Prospective cohort study with electronic medical record (EMR) surveillance for 7 years postpartum (August 2018–March 2026). Adjusted accelerated time-to-failure models estimated mLE8 associations with incident conditions.

**Setting:** A population-based prenatal cohort recruited from a large academic medical system in South Carolina.

**Participants:** Singleton pregnancies in individuals aged 18 to 44 years without pre-existing diabetes or cardiovascular disease (CVD)

**Exposures:** A 7-component mLE8 score assessed during pregnancy, incorporating hypertensive disorders of pregnancy (HDP), 50-g glucose tolerance test results, pre-pregnancy body mass index, smoking status, sleep adequacy, diet quality, and physical activity. Scores ranged from 0 to 100, with higher scores indicating more favorable CVH.

**Main Outcomes and Measures:** Post-delivery incident cardiometabolic conditions captured through EMRs and classified as chronic hypertensive conditions, chronic metabolic conditions (e.g., dyslipidemia, impaired glucose regulation), and CVD (e.g. cardiac arrest, cardiomyopathy). Time to incident diagnosis was measured in days from delivery.

**Results:** Among 1,225 pregnancies (mean age, 25.0 [5.3] years), 499 incident cardiometabolic events occurred over a median follow-up of 6.2 (2.8) years. Each 10-point higher mLE8 score was associated with a longer time to incident diagnosis of chronic hypertensive conditions (time ratio [TR], 1.26; 95% CI, 1.11–1.42) and chronic metabolic conditions (TR, 1.20; 95% CI, 1.11–1.29). Healthier HDP, glucose, BMI, and sleep scores were most strongly associated with longer time to diagnosis of chronic metabolic disease. Results were robust to sensitivity analyses excluding individuals who developed gestational diabetes or HDP.

**Conclusions and Relevance:** In this racially diverse, low-income cohort study of 1,225 pregnancies, better CVH during pregnancy was associated with a longer time to incident post-delivery diagnosis of cardiometabolic conditions. Pregnancy-based CVH assessment may help identify individuals with elevated and emerging cardiometabolic risk who could benefit from early, targeted intervention and enhanced longitudinal surveillance.

**Key Points:** **Question**: Is cardiovascular health during pregnancy, assessed using a modified Life’s Essential 8 score (LE8), associated with time to incident cardiometabolic disease after pregnancy?

**Findings**: In a racially diverse prospective pregnancy cohort with 7 years of electronic medical record follow-up, healthier mLE8 scores were associated with a significantly longer time to diagnosis of chronic hypertension and metabolic conditions. Associations persisted after excluding individuals with gestational diabetes or hypertensive disorders of pregnancy.

**Meaning**: Pregnancy cardiovascular health assessment using LE8 may identify individuals with elevated risk and latent vulnerability, and support earlier intervention during a sensitive window for prevention of long-term cardiometabolic disease.

## Introduction

Pregnancy is increasingly recognized as a prognostic window for long-term cardiometabolic health.^1, 2^ The physiologic demands of gestation can unmask latent dysfunction and serve as a natural cardiometabolic stress test.^2^ In individuals with underlying risk, this strain may manifest as adverse pregnancy outcomes (APOs), including gestational diabetes mellitus (GDM) and hypertensive disorders of pregnancy (HDP), which are associated with later type 2 diabetes, chronic hypertension, and cardiovascular disease.^3–7^ Pregnancy and the postpartum period therefore represent important opportunities to identify cardiometabolic risk early, particularly given persistent racial disparities in maternal morbidity and mortality in the United States.^7–11^

Despite growing clinical awareness, cardiovascular risk stratification during and after pregnancy largely remains reactive and poorly calibrated for reproductive-aged women.^1^ Traditional risk calculators were developed in older populations and generally exclude pregnancy-related factors, reducing their utility in reproductive-aged women.^12, 13,14–18^ While some have attempted to adapt traditional tools for pregnancy by accounting for APOs, these modified models frequently only marginally improve risk calculation.^19, 20^ Consequently, individuals at elevated risk may be overlooked during a period when prevention and early intervention may be especially valuable.^21^

The American Heart Association’s (AHA) Life’s Essential 8 (LE8) framework may offer a more comprehensive approach to risk assessment in pregnancy. Revised in 2022, LE8 integrates behavioral and clinical domains to characterize cardiovascular health across a continuum rather than relying only on over disease or APOs.^22^ This may be particularly relevant in pregnancy, where APO-based approaches may miss broader subclinical cardiometabolic vulnerability. For instance, while 6-14% of pregnant individuals experience GDM and 8% experience HDP,^23, 24^ over 95% of U.S. pregnant people do not meet criteria for high cardiovascular health (CVH) based on National Health and Nutrition Examination Data utilizing an early version of the LE8.^25^

However, the role of LE8 during pregnancy in identifying individuals at risk of later maternal cardiometabolic disease remains underexplored.^1^ Existing studies in pregnancy cohorts have largely been cross-sectional, and prospective work has focused primarily on neonatal outcomes.^26,27^ Although a recent UK Biobank found that higher LE8 scores attenuated CVD risk among women with prior APOs,^28^ LE8 was not measured during when the unique physiologic changes may reveal a population with emerging risk.^29^ Thus, its utility as a proactive risk stratification tool for long-term maternal cardiometabolic health remains unclear.

In this racially diverse, low-income cohort from South Carolina (SC), we examined the association between a modified LE8 (mLE8) score measured during pregnancy and incident cardiometabolic disease over seven years of follow-up. We hypothesized that higher mLE8 scores would be associated with a longer time to incident cardiometabolic disease after delivery.

## Materials and Methods

### Design, Setting, and Data

The Psychosocial Intervention and Inflammation in Centering (PIINC) Study (1R01HD092446–01A1, ClinicalTrials.gov: NCT04097548) enrolled participants from the Cradle randomized trial of group prenatal care (R01HD082311; NCT02640638) conducted in Greenville, SC.^30, 31^ All Cradle participants who had not yet delivered when PIINC began were eligible (N=1375). Individuals aged 14– 45 years with a singleton pregnancy were eligible for the parent trial stud; those with major medical comorbidities or conditions precluding group prenatal care were excluded. Full eligibility criteria have been reported elsewhere.^30^

PIINC began in August 2018 to collect biospecimens from participants and the last delivery occurred in October 2020. We excluded 31 participants with no encounters after delivery (**Supplemental Material 1**). Demographic, lifestyle, and stress surveys were completed at enrollment (≤20 weeks) and during the third trimester. Self-reported race and ethnicity, education, income, and insurance coverage during the prior 12 months were obtained from surveys; age at enrollment and gravidity were abstracted from electronic medical records (EMR) A second-trimester blood draw was obtained from PIINC participants. Pregnancy and delivery data were abstracted from the EMR (Epic Systems Corporation, Verona, WI). The PIINC study was approved by the NorthShore University HealthSystem Institutional Review Board (EH17–256), and all participants provided written informed consent. This study followed STROBE reporting guidelines.

### Primary Predictor, Cardiovascular Health During Pregnancy

CVH during pregnancy was assessed using the AHA LE8, which includes diet, physical activity, nicotine exposure, sleep health, BMI, non–high-density lipoprotein cholesterol (non-HDL-C), blood pressure, and blood glucose.^32^ Extensive details on the LE8 scoring can be found in Lloyd-Jones et al. 2022.^32^ Because non-HDL-C was unavailable in most participants (n=871), we used a modified LE8 (mLE8) score that excluded this component **(Supplemental Material 2**).

During the third trimester, surveys assessed diet, physical activity, and sleep. Diet quality was derived from a semi-quantitative food frequency questionnaire and adequacy/moderation of intake evaluated based on USDA recommendations (**Supplemental Material 3**).^33^ Physical activity was assessed using the International Physical Activity Questionnaire,^34–36^ and sleep using a single item on perceived sleep adequacy from the Health Promoting Lifestyle Profile II.^35, 36^ Smoking status was self-reported at enrollment. BMI was calculated from height and weight measured at the first antenatal visit (median 11.0 weeks’ gestation; [IQR]: 9.0–14.0). Glucose regulation was characterized using both a diagnosis of GDM based on Carpenter-Coustan criteria^37^ and plasma glucose levels obtained at 24-28 weeks following a 50-g oral glucose challenge test. The blood pressure component was scored on HDP abstracted from EMR after delivery.^38^ Thus, the mLE8 was conceptualized as a composite pregnancy cardiometabolic phenotype incorporating both underlying risk factors and clinical complications in a single pregnancy.

Each LE8 component was scored from 0-100, with higher scores indicating better CVH, and the total mLE8 score was calculated as the mean of available components.^39^ Scores were categorized as Low (0-49), Moderate (50-79) and High (80-100).^32^ In a subsample (n=354), second-trimester serum non-HDL cholesterol was quantified using targeted nuclear magnetic resonance spectroscopy (Nightingale Health©, Helsinki, Finland),^40, 41^ and only used in sensitivity analyses.

### Follow-Up and Outcome Ascertainment

Follow-up began seven days following delivery from the index pregnancy. For participants with a spontaneous abortion (n=36) or those missing delivery date (n=2), follow-up time began on their estimated delivery date using the last menstrual period or ultrasound dating. The surveillance concluded on March 20, 2026. Follow-up time was calculated from the start date to the first event. For each outcome, participants who remained event-free were censored at their last documented clinical encounter so that person-time reflected active observation within the health system.

Cardiometabolic conditions occurring outside a pregnancy episode were grouped as chronic metabolic conditions (e.g., diabetes, dyslipidemia), chronic hypertensive conditions (e.g., primary and secondary hypertension), and cardiovascular disease (e.g. cardiac arrest, cardiomyopathy, and ischemic heart disease). Outcome groups were defined using ICD-10 codes selected from the Elixhauser Comorbidity Index and prior literature (**Supplemental Material 4**).^42, 43^ Participants with chronic hypertension at enrollment or during the index pregnancy were excluded from the hypertensive outcome risk set (excluded n=129). Diagnosis of other cardiometabolic outcomes were not treated as censoring events; participants remained under observation for each outcome until event or censoring.

### Statistical Analysis

We estimated time ratios (TRs) and 95% CIs using accelerated failure time (AFT) models. Unlike hazard ratios, which measures instantaneous risk, AFT models directly parameterize differences in the timing of diagnosis. A TR>1.0 indicates a relative delay in diagnosis (longer time to event), while a TR<1.0 indicates an earlier diagnosis (accelerated onset). Weibull models were used for chronic hypertensin and metabolic outcomes and log-normal models for CVD because of sparse events and non-monotonic risk profiles.^44^ Model fit was assessed graphically using residual diagnostics. We assumed noninformative censoring, such that the probability of being censored (e.g., exiting the health system) was independent of the likelihood of developing a cardiometabolic condition, conditional on measured covariates. mLE8 was analyzed continuously per 10-point increase and categorically, with Low mLE8 as the referent.

#### Covariates

Model one adjusted for maternal age, model two additionally adjusted for ethnicity (Hispanic vs. non-Hispanic), gravidity, education, income, insurance, and attendance at a 6-week postpartum visit as a proxy for healthcare engagement. Race/ethnicity were included to account for confounding related to social and structural determinants of health.^45^ We did not adjust for delivery outcomes (e.g., preterm birth) because these may lie on the causal pathway between pregnancy health and later outcome.^46, 47^ P-values less than 0.05 were considered statistically significant. Analyses were performed in R statistical software version 4.4.0 (R Project for Statistical Computing).

#### Sensitivity analyses

We performed three sensitivity analyses. First, we repeated analyses after excluding women with GDM or HDP to assess whether associations were present independent of overt pregnancy complications. Second, in the subsample with non-HDL-C data (n=354), we repeated analyses including this component in the mLE8 score. Third, we adjusted for gestational age at enrollment to account for differences in early pregnancy weight gain.

## Results

The mean (SD) age at enrollment was 25.0 (5.3) years. The cohort was racially and ethnically diverse with 37.4% of participants identifying as non-Hispanic Black, 39.4% as White, and 22.1% as Hispanic. The sample was predominantly low-income: 39.3% reported an annual income of less than $20,000, whereas 3.0% $50,000 or more. Most participants (79.5%) had a high school education or less, and 66.2% had two or more pregnancies (**Table 1**). Gravidity and preterm birth differed across LE8 categories. The mean mLE8 score was in the Moderate range (69.9 [14.5]), as were most of the individual mLE8 components (**Figure 1**). The general pattern of Low, Moderate, and High mLE8 scores was similar across the different outcome-specific samples.

**Figure 1.**
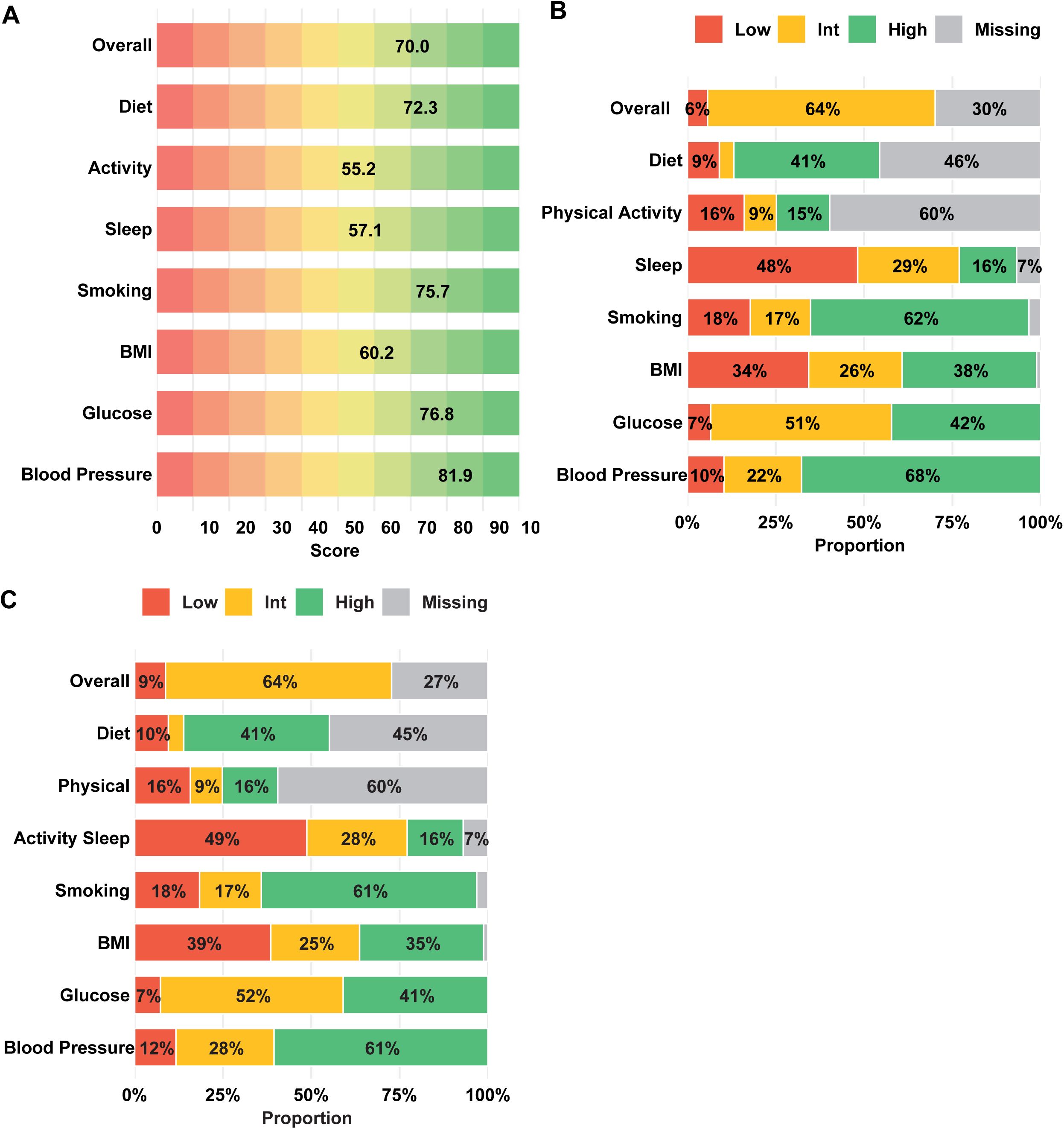
Mean Modified Life’s Essential Eight Total and Component Score Green = mLE8 80-100, orange = mLE8 50-79, red = mLE8 0-49 **A.** The mean modified Life’s Essential Eight (mLE8) total and component score assessed during pregnancy among 1,225 participants enrolled in the PIINC study. Bold numbers indicate the mean score for each component in the study sample. Blood lipids were only measured in a subsample of 324 women. **B.** Sample for analysis of incident hypertension (n=1096). The number of women with low, moderate, and high mLE8 scores for each component. **C.** Sample for analysis of incident CVD and metabolic outcomes, which represents the full analytic sample (n=1225). The number of women with low, moderate, and high mLE8 scores for each component.

**Table 1.**
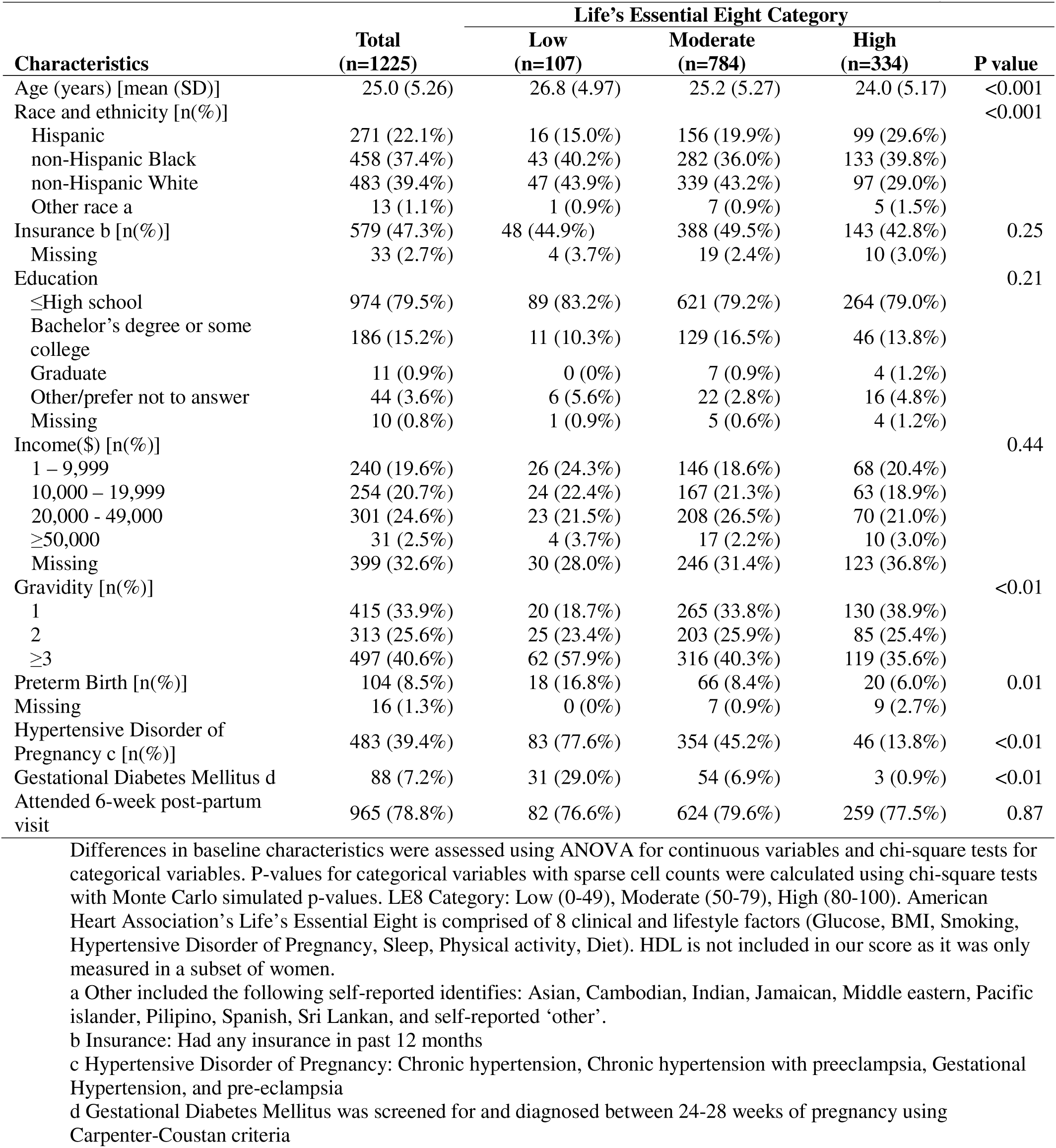
PIINC study descriptive characteristics and metrics of Life’s Essential Eight

| Characteristics | Life's Essential Eight Category |  |  |  | P value |
| --- | --- | --- | --- | --- | --- |
|  | Total<br>(n=1225) | Low<br>(n=107) | Moderate<br>(n=784) | High<br>(n=334) |  |
| Age (years) [mean (SD)] | 25.0 (5.26) | 26.8 (4.97) | 25.2 (5.27) | 24.0 (5.17) | <0.001 |
| Race and ethnicity [n(%)] |  |  |  |  | <0.001 |
| Hispanic | 271 (22.1%) | 16 (15.0%) | 156 (19.9%) | 99 (29.6%) |  |
| non-Hispanic Black | 458 (37.4%) | 43 (40.2%) | 282 (36.0%) | 133 (39.8%) |  |
| non-Hispanic White | 483 (39.4%) | 47 (43.9%) | 339 (43.2%) | 97 (29.0%) |  |
| Other race a | 13 (1.1%) | 1 (0.9%) | 7 (0.9%) | 5 (1.5%) |  |
| Insurance b [n(%)] | 579 (47.3%) | 48 (44.9%) | 388 (49.5%) | 143 (42.8%) | 0.25 |
| Missing | 33 (2.7%) | 4 (3.7%) | 19 (2.4%) | 10 (3.0%) |  |
| Education |  |  |  |  | 0.21 |
| ≤High school | 974 (79.5%) | 89 (83.2%) | 621 (79.2%) | 264 (79.0%) |  |
| Bachelor's degree or some college | 186 (15.2%) | 11 (10.3%) | 129 (16.5%) | 46 (13.8%) |  |
| Graduate | 11 (0.9%) | 0 (0%) | 7 (0.9%) | 4 (1.2%) |  |
| Other/prefer not to answer | 44 (3.6%) | 6 (5.6%) | 22 (2.8%) | 16 (4.8%) |  |
| Missing | 10 (0.8%) | 1 (0.9%) | 5 (0.6%) | 4 (1.2%) |  |
| Income(\$)[n(%)] | | | | | 0.44 |
| 1 – 9,999 | 240 (19.6%) | 26 (24.3%) | 146 (18.6%) | 68 (20.4%) |  |
| 10,000 – 19,999 | 254 (20.7%) | 24 (22.4%) | 167 (21.3%) | 63 (18.9%) |  |
| 20,000 - 49,000 | 301 (24.6%) | 23 (21.5%) | 208 (26.5%) | 70 (21.0%) |  |
| ≥50,000 | 31 (2.5%) | 4 (3.7%) | 17 (2.2%) | 10 (3.0%) |  |
| Missing | 399 (32.6%) | 30 (28.0%) | 246 (31.4%) | 123 (36.8%) |  |
| Gravidity [n(%)] |  |  |  |  | <0.01 |
| 1 | 415 (33.9%) | 20 (18.7%) | 265 (33.8%) | 130 (38.9%) |  |
| 2 | 313 (25.6%) | 25 (23.4%) | 203 (25.9%) | 85 (25.4%) |  |
| ≥3 | 497 (40.6%) | 62 (57.9%) | 316 (40.3%) | 119 (35.6%) |  |
| Preterm Birth [n(%)] | 104 (8.5%) | 18 (16.8%) | 66 (8.4%) | 20 (6.0%) | 0.01 |
| Missing | 16 (1.3%) | 0 (0%) | 7 (0.9%) | 9 (2.7%) |  |
| Hypertensive Disorder of Pregnancy c [n(%)] | 483 (39.4%) | 83 (77.6%) | 354 (45.2%) | 46 (13.8%) | <0.01 |
| Gestational Diabetes Mellitus d | 88 (7.2%) | 31 (29.0%) | 54 (6.9%) | 3 (0.9%) | <0.01 |
| Attended 6-week post-partum visit | 965 (78.8%) | 82 (76.6%) | 624 (79.6%) | 259 (77.5%) | 0.87 |

Differences in baseline characteristics were assessed using ANOVA for continuous variables and chi-square tests for categorical variables. P-values for categorical variables with sparse cell counts were calculated using chi-square tests with Monte Carlo simulated p-values. LE8 Category: Low (0-49), Moderate (50-79), High (80-100). American Heart Association's Life's Essential Eight is comprised of 8 clinical and lifestyle factors (Glucose, BMI, Smoking, Hypertensive Disorder of Pregnancy, Sleep, Physical activity, Diet). HDL is not included in our score as it was only measured in a subset of women.
a Other included the following self-reported identifies: Asian, Cambodian, Indian, Jamaican, Middle eastern, Pacific islander, Pilipino, Spanish, Sri Lankan, and self-reported 'other'.
b Insurance: Had any insurance in past 12 months
c Hypertensive Disorder of Pregnancy: Chronic hypertension, Chronic hypertension with preeclampsia, Gestational Hypertension, and pre-eclampsia
d Gestational Diabetes Mellitus was screened for and diagnosed between 24-28 weeks of pregnancy using Carpenter-Coustan criteria

The median (IQR) follow-up time was 6.2 (2.8) years, and mLE8 scores were not associated with censoring (p-value 0.15). The frequency of incident events, numbers at risk, and unadjusted Kaplan–Meier survival curves are shown in **Figure 2**. The most frequent event was chronic metabolic disease (n=257 [21%], e.g. diabetes, dyslipidemia), followed by chronic hypertensive conditions (n=209 [19%], e.g., primary and secondary hypertension), and cardiovascular disease (n=33 [3%], e.g., cardiac arrest, cardiomyopathy, and ischemic heart disease). The median (IQR) time to event for each outcome was 3.8 (3.5) years for chronic hypertension, 3.7 (3.6) years for chronic metabolic conditions, and 3.6 (4.0) years for CVD. The Kaplan-Meier curves showed longer event-free time for chronic hypertensive and metabolic conditions with higher mLE8 scores. These patterns persisted after excluding women with GDM or HDP (**Figure 3**). CVD curves showed limited separation, consistent with the low number of CVD events.

**Figure 2.**
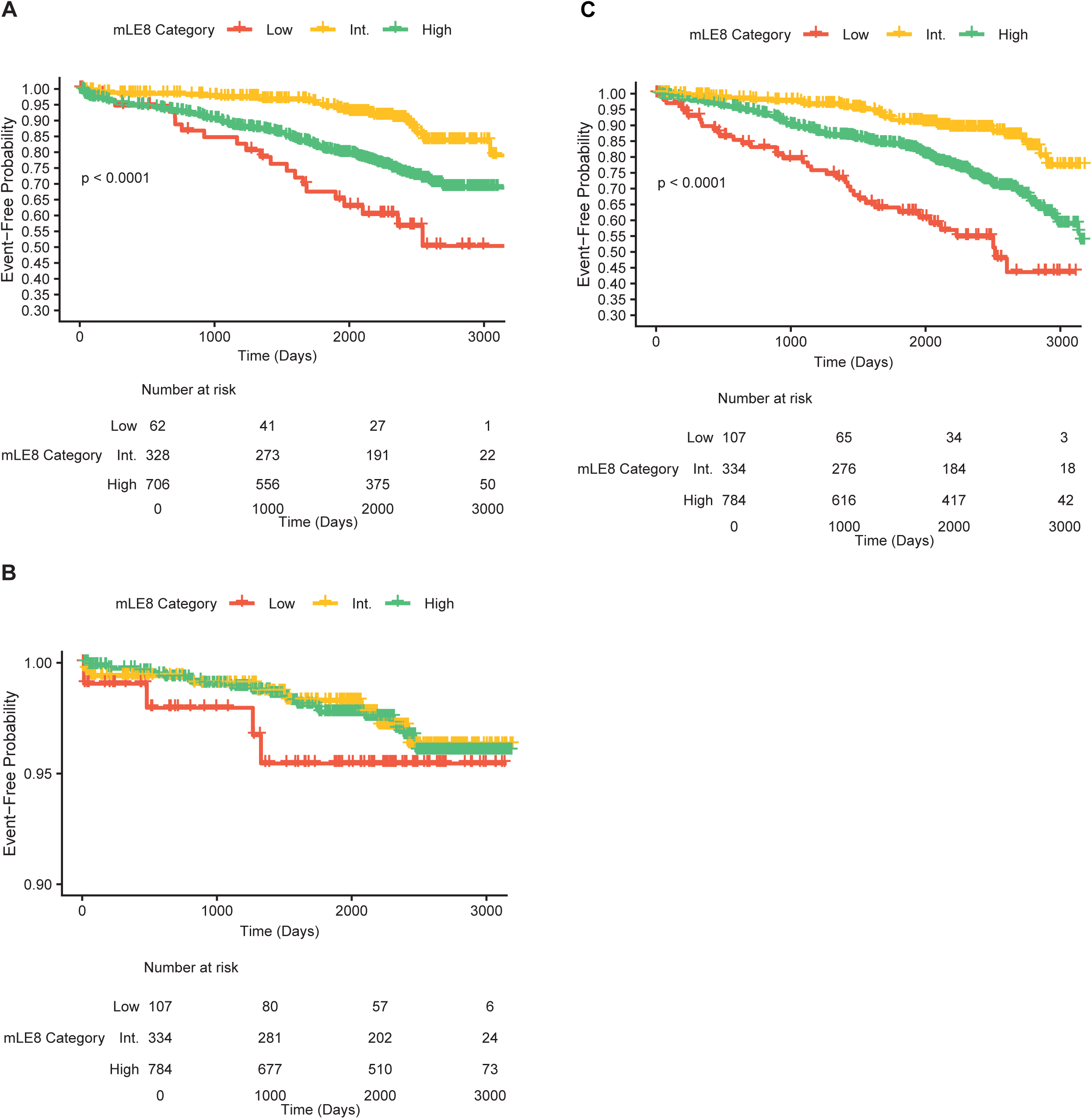
Unadjusted Kaplan–Meier Curves for Incident Cardiometabolic Outcomes in the Full Cohort by Modified Life’s Essential 8 (mLE8) Category Kaplan–Meier survival curves showing unadjusted time to incident chronic hypertensive conditions, chronic metabolic conditions, and cardiovascular disease (CVD) by modified Life’s Essential 8 (mLE8) score category. Green lines represent high CVH (mLE8 score 80–100), orange lines represent moderate CVH (mLE8 score 50–79), and red lines represent low CVH (mLE8 score 0–49). P values correspond to log-rank tests. A. Full cohort: incident chronic hypertensive conditions (n=1096). B. Full cohort: incident chronic metabolic conditions (n=1225). C. Full cohort: incident cardiovascular disease (n=1225).

**Figure 3.**
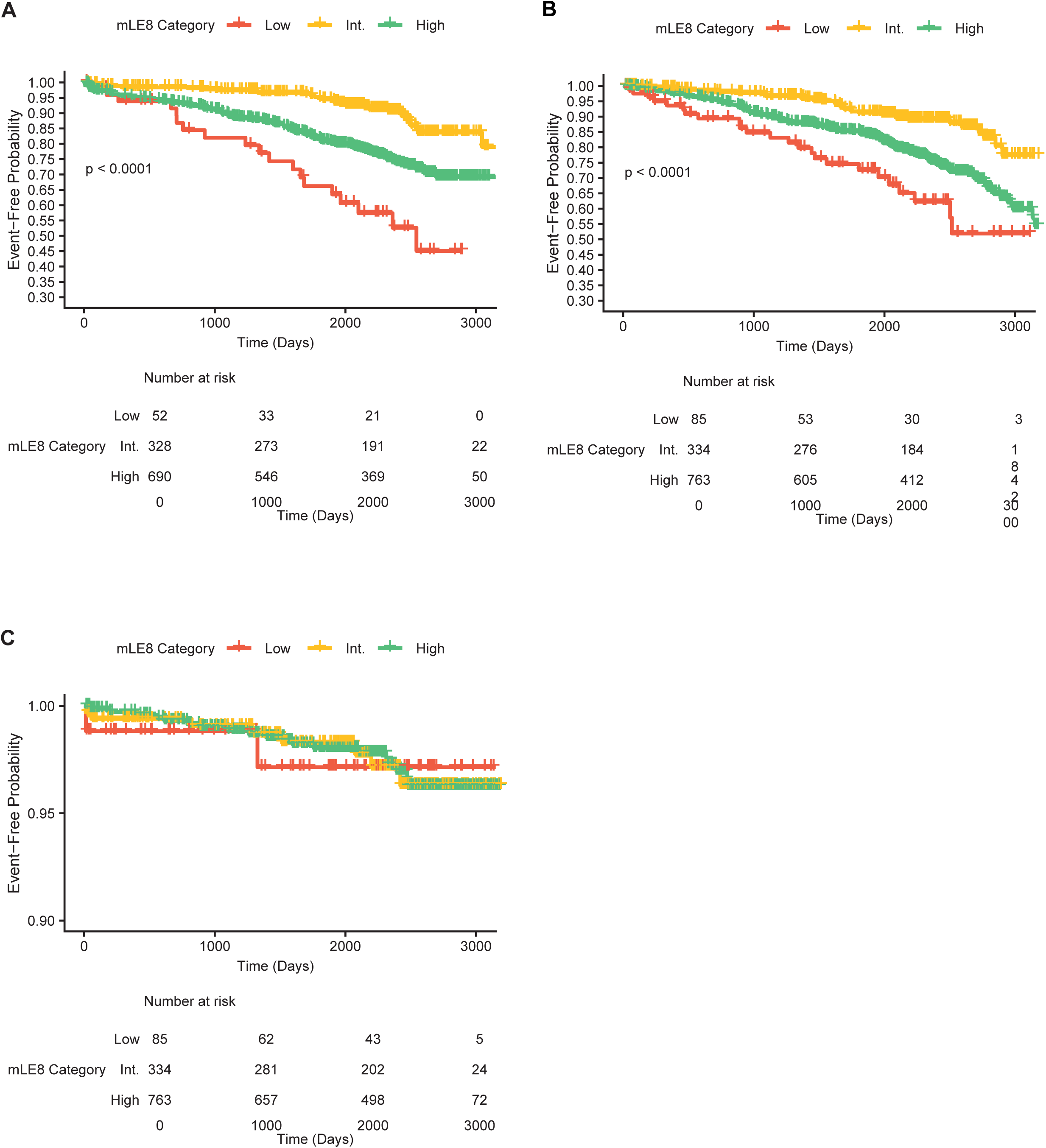
Unadjusted Kaplan–Meier Curves for Incident Cardiometabolic Outcomes in the Restricted Cohort by Modified Life’s Essential 8 (mLE8) Category Kaplan–Meier survival curves showing unadjusted time to incident chronic hypertensive conditions, chronic metabolic conditions, and cardiovascular disease (CVD) by modified Life’s Essential 8 (mLE8) score category. Green lines represent high CVH (mLE8 score 80–100), orange lines represent moderate CVH (mLE8 score 50–79), and red lines represent low CVH (mLE8 score 0–49). P values correspond to log-rank tests. B. Restricted cohort (excluding GDM or HDP): incident chronic hypertensive conditions (1070). C. Restricted cohort (excluding GDM or HDP): incident chronic metabolic conditions (n=1182). D. Restricted cohort (excluding GDM or HDP): incident cardiovascular disease (n=1182).

In adjusted models, each 10-unit increase in the mLE8 score was associated with a 26% longer time to diagnosis of chronic hypertensive conditions and a 28% longer time to diagnosis of chronic metabolic conditions (**Table 2**). Comparisons of high vs low mLE8 categories showed associations in the same direction but with wider confidence intervals. Moderate vs low mLE8 categories showed similar but smaller delays in onset. Healthier BMI, glucose, blood pressure, and sleep component scores were also associated with longer time to chronic hypertensive and metabolic conditions. Most findings were similar in magnitude and significance after excluding women with GDM or HDP. Higher mLE8 scores were also associated with longer time to CVD, although estimates were imprecise (**Supplemental Material 5**).

**Table 2.** Association of Life’s Essential 8 (LE8) and Time to Cardiometabolic Event

|  | Model 1 |  | Model 2 |  | Model 2.1 |  |
| --- | --- | --- | --- | --- | --- | --- |
|  | Time Ratio | P-value | Time Ratio | P-value | Time Ratio | P-value |
| **Total mLE8 score** |  |  |  |  |  |  |
| *Chronic hypertensive conditions<sup>a</sup>* |  |  |  |  |  |  |
| Per 10-unit increase | 1.34 (1.21, 1.48) | <0.01 | 1.26 (1.11, 1.42) | <0.01 | 1.28 (1.12, 1.46) | <0.01 |
| High vs. low | 4.13 (2.34, 7.28) | <0.01 | 3.89 (2.02, 7.50) | <0.01 | 4.32 (2.19, 8.51) | <0.01 |
| Moderate vs. low | 1.72 (1.10, 2.70) | 0.02 | 1.82 (1.10, 3.00) | 0.02 | 2.08 (1.23, 3.51) | 0.01 |
| *Chronic metabolic conditions<sup>b</sup>* |  |  |  |  |  |  |
| Per 10-unit increase | 1.25 (1.17, 1.33) | <0.01 | 1.20 (1.11, 1.29) | <0.01 | 1.14 (1.06, 1.23) | <0.01 |
| High vs. low | 4.13 (2.34, 7.28) | <0.01 | 3.89 (2.02, 7.50) | <0.01 | 4.32 (2.19, 8.51) | <0.01 |
| Moderate vs. low | 1.72 (1.10, 2.70) | 0.02 | 1.82 (1.10, 3.00) | 0.02 | 2.08 (1.23, 3.51) | 0.01 |
| **mLE8 Component score, per 10-unit increase** |  |  |  |  |  |  |
| *Chronic hypertensive conditions<sup>a</sup>* |  |  |  |  |  |  |
| <u>Behavioral Factors</u> |  |  |  |  |  |  |
| Diet | 0.99 (0.93, 1.05) | 0.75 | 0.98 (0.92, 1.05) | 0.62 | 0.99 (0.92, 1.06) | 0.70 |
| Physical activity | 1.04 (0.97, 1.11) | 0.31 | 1.07 (0.99, 1.17) | 0.10 | 1.06 (0.98, 1.16) | 0.15 |
| Sleep | 1.07 (1.01, 1.13) | 0.01 | 1.05 (0.98, 1.12) | 0.14 | 1.05 (0.98, 1.11) | 0.18 |
| Smoking | 1.01 (0.98, 1.05) | 0.46 | 0.97 (0.93, 1.01) | 0.18 | 0.98 (0.93, 1.02) | 0.26 |
| <u>Health Factors</u> |  |  |  |  |  |  |
| BMI | 1.07 (1.03, 1.11) | <0.01 | 1.08 (1.03, 1.13) | <0.01 | 1.08 (1.03, 1.13) | <0.01 |
| Glucose | 1.04 (0.97, 1.10) | 0.27 | 1.06 (0.98, 1.15) | 0.15 | 1.05 (0.96, 1.14) | 0.28 |
| Hypertension | 1.20 (1.14, 1.26) | <0.01 | 1.21 (1.14, 1.28) | <0.01 | 1.23 (1.15, 1.30) | <0.01 |
| *Chronic metabolic conditions<sup>b</sup>* |  |  |  |  |  |  |
| <u>Behavioral Factors</u> |  |  |  |  |  |  |
| Diet | 1.00 (0.96, 1.04) | 0.97 | 1.00 (0.95, 1.05) | 0.97 | 1.00 (0.96, 1.05) | 0.95 |
| Physical activity | 0.99 (0.93, 1.04) | 0.62 | 1.00 (0.93, 1.07) | 0.96 | 1.00 (0.94, 1.06) | 0.99 |
| Sleep | 1.05 (1.01, 1.09) | 0.01 | 1.05 (1.00, 1.09) | 0.03 | 1.04 (1.00, 1.08) | 0.04 |
| Smoking | 0.99 (0.97, 1.02) | 0.46 | 0.98 (0.96, 1.01) | 0.30 | 0.99 (0.96, 1.02) | 0.44 |
| <u>Health Factors</u> |  |  |  |  |  |  |
| BMI | 1.10 (1.07, 1.13) | <0.01 | 1.07 (1.04, 1.11) | <0.01 | 1.05 (1.02, 1.08) | <0.01 |
| Glucose | 1.18 (1.13, 1.23) | <0.01 | 1.15 (1.10, 1.21) | <0.01 | 1.11 (1.05, 1.17) | <0.01 |
| Hypertension | 1.06 (1.03, 1.10) | <0.01 | 1.06 (1.03, 1.10) | <0.01 | 1.04 (1.00, 1.07) | 0.07 |

a includes all women without chronic hypertension prior to pregnancy n=1017 (e.g., excludes Hypertensive Disorders of Pregnancy: chronic hypertension without preeclampsia [n=92] and chronic hypertension superimposed on preeclampsia [n=30]).
b includes all women n=1256. Outcomes include Diabetes, Dyslipidemia, Metabolic disorders, Metabolic syndrome, Prediabetes/Hyperglycemia
Model 1: adjusted for maternal age
Model 2: adjusted for maternal age, ethnicity (Hispanic vs. non-Hispanic), gravidity (continuously), education, income, insurance.
Model 2.1: Excludes women with GDM and HDP. Adjusted for maternal ethnicity (Hispanic vs. non-Hispanic), gravidity (continuously), education, income, insurance.

In sensitivity analyses, inclusion of the non-HDL score and additional adjustment for gestational age at enrollment produced findings similar in direction, magnitude, and significance to the fully adjusted models.

## Discussion

In this racially and ethnically diverse longitudinal pregnancy cohort, more favorable mLE8 during pregnancy was associated with longer time to incident cardiometabolic and vascular disorders over seven years of EMR surveillance after delivery. Healthier scores on the glucose, BMI, hypertension, and sleep components were associated with delayed onset of chronic hypertensive and metabolic conditions. Healthier hypertension component scores were also associated with a delayed onset of cardiovascular disease, although estimates for cardiovascular disease were imprecise because of low event counts. These findings replicate prior work focused on APOs and later cardiometabolic risk,^5, 6, 48–50^ and highlight that the critical but narrow focus on APOs may overlook broader subclinical vulnerability relevant for maternal health in the decade following pregnancy.

Several pathways may explain why healthier LE8 scores during pregnancy were associated with a longer time to later cardiometabolic diagnosis. Indeed, healthier LE8 scores in pregnancy may capture both better preconception CVH and a more adaptive response to the cardiometabolic stress of pregnancy. Our findings are generally consistent with prior work linking poorer CVH before or during pregnancy with later maternal risk.^14, 51–53^ For instance, among the Generation R study (Netherlands), worse CVH assessed using an earlier version of LE8 (Life’s Simple Seven) was associated with greater carotid intima media thickness two decades later.^51^ In the U.S., pre-pregnancy CVH in the CARDIA study predicted GDM, HDP, and coronary artery calcium; and mediation analyses showed that APOs explained only a small portion of the association between pre-pregnancy CVH and later disease.^52, 53^ Consistent with that literature, associations in our study were largely unchanged after excluding women with GDM or HDP, supporting the idea that LE8 may identify vulnerability beyond overt pregnancy complications. Because these conditions develop sequentially and share underlying pathophysiology (e.g., hypertension preceding cardiovascular disease), our estimates should be interpreted as condition-specific rather than independent effects across outcomes.

Use of the LE8 framework also highlighted specific components most strongly associated with later health. Lower BMI and blood pressure, and improved glucose regulation were consistently linked to delayed onset of chronic metabolic and hypertensive conditions, in line with extensive evidence linking these domains to long-term cardiometabolic risk.^32, 46^ Sleep showed a somewhat weak association with cardiometabolic risk, which aligns somewhat with the literature indicating that sleep is associated with both APOs and cardiometabolic risk. ^9, 54–56^ We did not observe associations for smoking, physical activity or diet. These null findings should be interpreted cautiously given the low prevalence of smoking, possible exposure misclassification, reliance on brief self-reported measures, and the relatively young cohort with limited numbers of some later vascular outcomes. Despite this, there is strong evidence for the importance of these behaviors in preventing the development of CVD and its risk factors.^57^ Future research evaluating the implementation of physical activity and nutrition assessment in clinical practice is needed to determine the most appropriate tools for translation.

### Implications for Pregnancy as a CVH Assessment Window

Pregnancy may be a uniquely informative window for CVH assessment because the physiologic demands of gestation can reveal latent metabolic and vascular dysfunction.^29^ In this context, LE8 offers a pragmatic multidomain framework that could help identify individuals with emerging vulnerability who remain below conventional diagnostic thresholds. This may be particularly relevant for reproductive age women who otherwise have limited interaction with the healthcare system.^58, 59^ Because most LE8 components are already assessed during pregnancy, the utilization of the LE8 framework for postpartum risk stratification may be feasible. These findings also align with broader efforts to improve recognition of maternal cardiovascular risk in obstetric care, such as the use of maternal safety bundles.^10, 60^ Pregnancy-based CVH assessment may complement these efforts by helping identify a broader population with emerging metabolic and vascular vulnerability, beyond those already recognized as highest risk.

In turn, this could support earlier transition to primary care and more tailored prevention through enhanced postpartum monitoring of blood pressure, weight, and glucose, targeted lifestyle counseling, and referral of higher-risk patients for co-management or specialty follow-up when appropriate, or other supportive services. Such risk stratification may also inform interconception care, identify women at elevated risk for complications in subsequent pregnancies, and support earlier action before midlife and menopause, when cardiometabolic risk trajectories may already be established.^61^ At the same time, pregnancy-specific adaptation of LE8 requires further validation, particularly because some metabolic markers change dynamically across gestation and optimal pregnancy-specific thresholds remain uncertain. Future studies are needed to refine pregnancy-specific LE8 adaptations and to determine how coordinated care across obstetrics, primary care, cardiology, and community-based supports (e.g., doula care, home visiting programs, patient navigators or other community health programming) may reduce cardiovascular risk during, between, and after pregnancies These findings may be particularly relevant in populations facing social and structural barriers to preventative care. Socioeconomically marginalized and racially and ethnically underrepresented populations are more likely to enter pregnancy with poor CVH,^14, 25, 57, 62, 63^ experience APOs,^56^ and face barriers to follow-up care after delivery. ^64–66^ Unmet social and structural needs before and during pregnancy, including barriers related to poverty, housing, food access, insurance coverage, chronic stress, and limited access to preventive care, may themselves contribute to poor long-term CVH.^14, 63^ As such, pregnancy-based CVH assessment may therefore represent not only a clinical opportunity but also an equity-relevant strategy for identifying risk during a period when women have greater healthcare engagement.^58^ However, equitable implementation will be essential so that pregnancy-based risk stratification does not further widen disparities in settings with limited prenatal care infrastructure or maternity care access.

### Strengths and Limitations

This study has several strengths and limitations. Strengths include longitudinal EMR surveillance in a large cohort of pregnant women from a population often underrepresented in long-term cardiometabolic research. However, EMR diagnosis dates likely reflect time to clinical recognition rather than true biological onset, and differences in healthcare utilization may influence that timing. Consequently, our Time Ratios should be interpreted as the acceleration or deceleration of clinical diagnosis rather than biological onset. Although disease onset and clinical diagnosis may not occur simultaneously, in this young, marginalized population, the time to clinical recognition is a critical public health metric, as it represents the point at which medical intervention and secondary prevention begin. Our reliance on last encounter date for censoring reduces person-time misclassification but may underestimate survival time among participants who remained disease-free after their last encounter. Because follow-up ended on March 20, 2026, participants who delivered later in the study window contributed less maximum observable follow-up time, which may have reduced capture of later-onset outcomes. Misclassification of self-reported diet, physical activity, and smoking is possible and thus the null associations related to some of these components should be interpreted cautiously.^67, 68^ We adapted the LE8 scoring algorithm to incorporate pregnancy complications; while this may increase the relevance of LE8 to pregnant populations, additional validation is needed. Relatedly, the blood pressure component was based on the presence of HDP rather than continuous blood pressure values, potentially missing more nuanced associations. A unique strength of our study was the inclusion of large sample of women from a population that often experiences structural marginalization, extending CVH research to a group often systematically excluded from longitudinal cardiometabolic studies. However, as such, our findings may not generalize to populations with different sociodemographic characteristics or different baseline CVH. Studies are needed to validate the mLE8 score in other populations, and future areas for research could incorporate subsequent pregnancy complications which are associated with greater morbidity and mortality.^9, 69^ Importantly, future studies with inclusive recruitment will be essential to ensure that future CVH research advances both scientific rigor and health equity. Without adequate representation of these groups, pregnancy-based CVH tools and risk stratification approaches may be less equitable, less generalizable, and less likely to benefit the populations most affected by cardiometabolic inequities.

## Conclusions

In this longitudinal pregnancy cohort study with seven years of EMR surveillance, better CVH as assessed by a mLE8 during pregnancy was associated with delayed time to diagnosis of chronic hypertension, metabolic disorders, and, less precisely, cardiovascular disease during the years following delivery. These associations persisted after excluding women with GDM or HDP, suggesting pregnancy CVH assessment may identify subclinical vulnerability beyond overt complications. Further research is needed to validate pregnancy-adapted LE8 measures and determine how they can be incorporated into maternal risk stratification and prevention frameworks.

## Data Availability

All data produced in the present study are available upon reasonable request to the authors

## Declarations

### Ethics approval and consent to participate

The PIINC study (ClinicalTrials.gov: NCT04097548) was approved by the NorthShore University HealthSystem Institutional Review Board (EH17–256). All participants gave written informed consent at enrollment.

### Availability of data and materials

Data are not available publicly due to participant consent; however, reasonable requests can be made to the PIINC study team via the corresponding author.

## Competing interests

The authors declare that they have no competing interests

## Authors’ contributions

ECF conceptualized the study, analyzed the data, and drafted the manuscript. SP conducted independent data analysis to verify findings in SAS. SP and AP supported literature review. AHC oversaw sample collection and patient care of PIINC participants. SR provided critical feedback on dietary and physical activity score. AF, LKD, ESB, LME, AB, GM, SR, AHC edited the manuscript, supported the interpretation of results, and provided critical review of the final version.

## Acknowledgements

We sincerely thank the CRADLE/PIINC participants, study staff, and past investigators for their valuable contributions to this study and mentorship. We acknowledge the data management and harmonization efforts undertaken by Dr. Jessie Britt, for without these the analyses would not be possible. Dr. Elizabeth Suarez provided valuable feedback during the revision, and we thank her for her insights on censoring approaches. The conceptualization of cardiovascular health was a multidisciplinary process that spanned several decades. We thank and acknowledge the American Heart Association and all those who contributed to the operationalization of cardiovascular health and the Life’s Essential Eight score.^32^

Dr. Ellen C. Francis had full access to all the data in the study and takes responsibility for the integrity of the data and the accuracy of the data analysis.

## Funding

This research was funded by the Eunice Kennedy Shriver National Institute of Child Health and Human Development grant numbers R00 HD108272, R01HD082311, R01HD092446; the National Heart Lung and Blood Institute grant number R25HL146166-05; and the National Institute of Environmental Health Sciences P30ES005022

## Role of the Funder/Sponsor

The funders had no role in the design and conduct of the study; collection, management, analysis, and interpretation of the data; preparation, review, or approval of the manuscript; and decision to submit the manuscript for publication.

## Notes

### Competing Interest Statement

The authors have declared no competing interest.

### Clinical Trial

NCT04097548, NCT02640638

### Author Declarations

The PIINC study was approved by the NorthShore University HealthSystem Institutional Review Board (EH17-256), and all participants provided written informed consent

## References

1. Lewey J, Beckie TM, Brown HL, Brown SD, Garovic VD, Khan SS, Miller EC, Sharma G, Mehta LS. Opportunities in the Postpartum Period to Reduce Cardiovascular Disease Risk After Adverse Pregnancy Outcomes: A Scientific Statement From the American Heart Association. Circulation. 2024;149(7):e330–e46. Epub 20240212. doi: 10.1161/cir.0000000000001212. PubMed PMID: 38346104; PMCID: PMC11185178.

2. Parikh NI, Gonzalez JM, Anderson CAM, Judd SE, Rexrode KM, Hlatky MA, Gunderson EP, Stuart JJ, Vaidya D. Adverse Pregnancy Outcomes and Cardiovascular Disease Risk: Unique Opportunities for Cardiovascular Disease Prevention in Women: A Scientific Statement From the American Heart Association. Circulation. 2021;143(18):e902–e16. Epub 20210329. doi: 10.1161/cir.0000000000000961. PubMed PMID: 33779213.

3. Ehrenthal DB, McNeil RB, Crenshaw EG, Bairey Merz CN, Grobman WA, Parker CB, Greenland P, Pemberton VL, Zee PC, Scifres CM, Polito L, Saade G. Adverse Pregnancy Outcomes and Future Metabolic Syndrome. Journal of Women’s Health. 2023;32(9):932–41. doi: 10.1089/jwh.2023.0026.

4. Hutchins F, El Khoudary SR, Catov J, Krafty R, Colvin A, Barinas-Mitchell E, Brooks MM. Excessive Gestational Weight Gain and Long-Term Maternal Cardiovascular Risk Profile: The Study of Women’s Health Across the Nation. J Womens Health (Larchmt). 2022;31(6):808–18. Epub 20220418. doi: 10.1089/jwh.2021.0449. PubMed PMID: 35442810; PMCID: PMC9245790.

5. Venkatesh KK, Khan SS, Yee LM, Wu J, McNeil R, Greenland P, Chung JH, Levine LD, Simhan HN, Catov J, Scifres C, Reddy UM, Pemberton VL, Saade G, Bairey Merz CN, Grobman WA. Adverse Pregnancy Outcomes and Predicted 30-Year Risk of Maternal Cardiovascular Disease 2-7 Years After Delivery. Obstet Gynecol. 2024;143(6):775–84. Epub 20240405. doi: 10.1097/aog.0000000000005569. PubMed PMID: 38574364; PMCID: PMC11098696.

6. Venkatesh KK, Grobman WA, Wu J, Shah NS, Pencina M, Costantine MM, Landon MB, Catalano P, Lowe WL, Scholtens DM, Khan SS. Hypertensive disorders of pregnancy and gestational diabetes mellitus and predicted risk of maternal cardiovascular disease 10-14 years after delivery: A prospective cohort. Diabet Med. 2025;42(5):e15516. Epub 20250117. doi: 10.1111/dme.15516. PubMed PMID: 39825470; PMCID: PMC12005981.

7. Xie W, Wang Y, Xiao S, Qiu L, Yu Y, Zhang Z. Association of gestational diabetes mellitus with overall and type specific cardiovascular and cerebrovascular diseases: systematic review and meta-analysis. Bmj. 2022;378:e070244. Epub 20220921. doi: 10.1136/bmj-2022-070244. PubMed PMID: 36130740; PMCID: PMC9490552.

8. Grandi SM, Filion KB, Yoon S, Ayele HT, Doyle CM, Hutcheon JA, Smith GN, Gore GC, Ray JG, Nerenberg K, Platt RW. Cardiovascular Disease-Related Morbidity and Mortality in Women With a History of Pregnancy Complications. Circulation. 2019;139(8):1069–79. doi: 10.1161/circulationaha.118.036748. PubMed PMID: 30779636.

9. Wang Y-X, Mitsunami M, Manson JE, Gaskins AJ, Rich-Edwards JW, Wang L, Zhang C, Chavarro JE. Association of Gestational Diabetes With Subsequent Long-Term Risk of Mortality. JAMA Internal Medicine. 2023;183(11):1204–13. doi: 10.1001/jamainternmed.2023.4401.

10. Brown HL, Warner JJ, Gianos E, Gulati M, Hill AJ, Hollier LM, Rosen SE, Rosser ML, Wenger NK, American Heart A, the American College of O, Gynecologists. Promoting Risk Identification and Reduction of Cardiovascular Disease in Women Through Collaboration With Obstetricians and Gynecologists: A Presidential Advisory From the American Heart Association and the American College of Obstetricians and Gynecologists. Circulation. 2018;137(24):e843–e52. Epub 20180510. doi: 10.1161/CIR.0000000000000582. PubMed PMID: 29748185.

11. Wang E, Glazer KB, Howell EA, Janevic TM. Social Determinants of Pregnancy-Related Mortality and Morbidity in the United States: A Systematic Review. Obstet Gynecol. 2020;135(4):896–915. PubMed PMID: 32168209.

12. Dam V, Onland-Moret NC, Verschuren WMM, Boer JMA, Benschop L, Franx A, Moons KGM, Boersma E, van der Schouw YT. Cardiovascular risk model performance in women with and without hypertensive disorders of pregnancy. Heart. 2019;105(4):330–6. doi: 10.1136/heartjnl-2018-313439.

13. Moe K, Sugulle M, Dechend R, Staff AC. Risk prediction of maternal cardiovascular disease one year after hypertensive pregnancy complications or gestational diabetes mellitus. Eur J Prev Cardiol. 2020;27(12):1273–83. Epub 20191010. doi: 10.1177/2047487319879791. PubMed PMID: 31600083.

14. Cameron NA, Huang X, Petito LC, Ning H, Shah NS, Yee LM, Perak AM, Haas DM, Mercer BM, Parry S, Saade GR, Silver RM, Simhan HN, Reddy UM, Varagic J, Licon E, Greenland P, Lloyd-Jones DM, Kershaw KN, Grobman WA, Khan SS. Determinants of Racial and Ethnic Differences in Maternal Cardiovascular Health in Early Pregnancy. Circ Cardiovasc Qual Outcomes. 2025;18(3):e011217. Epub 20250114. doi: 10.1161/circoutcomes.124.011217. PubMed PMID: 39807595; PMCID: PMC11919558.

15. Mora S, Wenger NK, Cook NR, Liu J, Howard BV, Limacher MC, Liu S, Margolis KL, Martin LW, Paynter NP, Ridker PM, Robinson JG, Rossouw JE, Safford MM, Manson JE. Evaluation of the Pooled Cohort Risk Equations for Cardiovascular Risk Prediction in a Multiethnic Cohort From the Women’s Health Initiative. JAMA Internal Medicine. 2018;178(9):1231–40. doi: 10.1001/jamainternmed.2018.2875.

16. Mehta LS, Velarde GP, Lewey J, Sharma G, Bond RM, Navas-Acien A, Fretts AM, Magwood GS, Yang E, Blumenthal RS, Brown RM, Mieres JH. Cardiovascular Disease Risk Factors in Women: The Impact of Race and Ethnicity: A Scientific Statement From the American Heart Association. Circulation. 2023;147(19):1471–87. Epub 20230410. doi: 10.1161/cir.0000000000001139. PubMed PMID: 37035919; PMCID: PMC11196122.

17. Rodriguez F, Chung S, Blum MR, Coulet A, Basu S, Palaniappan LP. Atherosclerotic Cardiovascular Disease Risk Prediction in Disaggregated Asian and Hispanic Subgroups Using Electronic Health Records. J Am Heart Assoc. 2019;8(14):e011874. Epub 20190711. doi: 10.1161/jaha.118.011874. PubMed PMID: 31291803; PMCID: PMC6662141.

18. Khan SS, Matsushita K, Sang Y, Ballew SH, Grams ME, Surapaneni A, Blaha MJ, Carson AP, Chang AR, Ciemins E, Go AS, Gutierrez OM, Hwang SJ, Jassal SK, Kovesdy CP, Lloyd-Jones DM, Shlipak MG, Palaniappan LP, Sperling L, Virani SS, Tuttle K, Neeland IJ, Chow SL, Rangaswami J, Pencina MJ, Ndumele CE, Coresh J. Development and Validation of the American Heart Association’s PREVENT Equations. Circulation. 2024;149(6):430–49. PubMed PMID: 37947085.

19. Markovitz AR, Stuart JJ, Horn J, Williams PL, Rimm EB, Missmer SA, Tanz LJ, Haug EB, Fraser A, Timpka S, Klykken B, Dalen H, Romundstad PR, Rich-Edwards JW, Åsvold BO. Does pregnancy complication history improve cardiovascular disease risk prediction? Findings from the HUNT study in Norway. Eur Heart J. 2019;40(14):1113–20. Epub 2019/01/01. doi: 10.1093/eurheartj/ehy863. PubMed PMID: 30596987; PMCID: PMC6451770.

20. Saei Ghare Naz M, Sheidaei A, Aflatounian A, Azizi F, Ramezani Tehrani F. Does Adding Adverse Pregnancy Outcomes Improve the Framingham Cardiovascular Risk Score in Women? Data from the Tehran Lipid and Glucose Study. J Am Heart Assoc. 2022;11(2):e022349. Epub 2022/01/13. doi: 10.1161/jaha.121.022349. PubMed PMID: 35016530; PMCID: PMC9238524.

21. Olds DL, Kitzman H, Knudtson MD, Anson E, Smith JA, Cole R. Effect of home visiting by nurses on maternal and child mortality: results of a 2-decade follow-up of a randomized clinical trial. JAMA Pediatr. 2014;168(9):800–6. PubMed PMID: 25003802.

22. Lloyd-Jones DM, Allen NB, Anderson CAM, Black T, Brewer LC, Foraker RE, Grandner MA, Lavretsky H, Perak AM, Sharma G, Rosamond W. Life’s Essential 8: Updating and Enhancing the American Heart Association’s Construct of Cardiovascular Health: A Presidential Advisory From the American Heart Association. Circulation. 2022;146(5):e18–e43. Epub 20220629. doi: 10.1161/cir.0000000000001078. PubMed PMID: 35766027; PMCID: PMC10503546.

23. Sweeting A, Hannah W, Backman H, Catalano P, Feghali M, Herman WH, Hivert MF, Immanuel J, Meek C, Oppermann ML, Nolan CJ, Ram U, Schmidt MI, Simmons D, Chivese T, Benhalima K. Epidemiology and management of gestational diabetes. Lancet. 2024;404(10448):175–92. Epub 20240620. doi: 10.1016/s0140-6736(24)00825-0. PubMed PMID: 38909620.

24. Bruno AM, Allshouse AA, Metz TD, Theilen LH. Trends in Hypertensive Disorders of Pregnancy in the United States From 1989 to 2020. Obstet Gynecol. 2022;140(1):83–6. Epub 20220607. doi: 10.1097/aog.0000000000004824. PubMed PMID: 35849460; PMCID: PMC10472797.

25. Perak AM, Ning H, Khan SS, Van Horn LV, Grobman WA, Lloyd-Jones DM. Cardiovascular Health Among Pregnant Women, Aged 20 to 44 Years, in the United States. J Am Heart Assoc. 2020;9(4):e015123. Epub 20200217. doi: 10.1161/jaha.119.015123. PubMed PMID: 32063122; PMCID: PMC7070227.

26. Khan SS, Brewer LC, Canobbio MM, Cipolla MJ, Grobman WA, Lewey J, Michos ED, Miller EC, Perak AM, Wei GS, Gooding H. Optimizing Prepregnancy Cardiovascular Health to Improve Outcomes in Pregnant and Postpartum Individuals and Offspring: A Scientific Statement From the American Heart Association. Circulation. 2023;147(7):e76–e91. Epub 20230213. doi: 10.1161/cir.0000000000001124. PubMed PMID: 36780391; PMCID: PMC10080475.

27. Jing G, Wei Q, Zou J, Zhang Y, Shi H, Gao X. Longitudinal association between maternal cardiovascular health in pregnancy and child birth outcomes. Sci Rep. 2024;14(1):15355. Epub 20240704. doi: 10.1038/s41598-024-66029-6. PubMed PMID: 38961151; PMCID: PMC11222450.

28. Chen H, Li R, Liu S, Zhao S, Guo T, Tian S, Zhong J, Tang Z, Ge Z, Xia J, Geng T, Pan X, Pan A, Qian F, Liu G. Life’s Essential 8 and cardiovascular disease in women with a history of adverse pregnancy outcomes. Eur J Prev Cardiol. 2025. Epub 20250131. doi: 10.1093/eurjpc/zwaf021. PubMed PMID: 39887026.

29. Rich-Edwards JW. Reproductive Health as a Sentinel of Chronic Disease in Women. Women’s Health. 2009;5(2):101–5. doi: 10.2217/17455057.5.2.101.

30. Chen L, Crockett AH, Covington-Kolb S, Heberlein E, Zhang L, Sun X. Centering and Racial Disparities (CRADLE study): rationale and design of a randomized controlled trial of centeringpregnancy and birth outcomes.

31. BMC Pregnancy Childbirth. 2017;17(1):118. Epub 2017/04/14. doi: 10.1186/s12884-017-1295-7. PubMed PMID: 28403832.

31. Crockett AH, Chen L, Heberlein EC, Britt JL, Covington-Kolb S, Witrick B, Doherty E, Zhang L, Borders A, Keenan-Devlin L, Smart B, Heo M. Group vs traditional prenatal care for improving racial equity in preterm birth and low birthweight: the Centering and Racial Disparities randomized clinical trial study. Am J Obstet Gynecol. 2022;227(6):893.e1–.e15. Epub 2022/09/17. doi: 10.1016/j.ajog.2022.06.066. PubMed PMID: 36113576; PMCID: PMC9729420.

32. Lloyd-Jones DM, Allen NB, Anderson CAM, Black T, Brewer LC, Foraker RE, Grandner MA, Lavretsky H, Perak AM, Sharma G, Rosamond W, on behalf of the American Heart A. Life’s Essential 8: Updating and Enhancing the American Heart Association’s Construct of Cardiovascular Health: A Presidential Advisory From the American Heart Association. Circulation. 2022;146(5):e18–e43. doi: 10.1161/CIR.0000000000001078.

33. United States Department of Agriculture Food and Nutrition Service. Women, Infants, and Children (WIC): United States Department of Agriculture; 2018 [cited 2018 May 25th]. Available from: https://www.fns.usda.gov/wic/women-infants-and-children-wic.

34. Craig CL, Marshall AL, Sjostrom M, Bauman AE, Booth ML, Ainsworth BE, Pratt M, Ekelund U, Yngve A, Sallis JF, Oja P. International physical activity questionnaire: 12-country reliability and validity. Medicine and science in sports and exercise. 2003;35(8):1381–95. Epub 2003/08/06. doi: 10.1249/01.MSS.0000078924.61453.FB. PubMed PMID: 12900694.

35. Walker SN, Kerr MJ, Pender NJ, Secrist KR. A Spanish Language Version Of the Health-Promoting Lifestyle Profile. Nursing Research. 1990;39(5):268–73.

36. Walker SN, Sechrist KR, Pender NJ. The health-promoting lifestyle profile: development and psychometric characteristics. Nursing research. 1987;36(2):76–81.

37. ACOG Committee on Obstetric Practice. ACOG Practice Bulletin No. 190: Gestational Diabetes Mellitus. Obstet Gynecol. 2018;131(2):e49–e64. Epub 2018/01/26. doi: 10.1097/aog.0000000000002501. PubMed PMID: 29370047.

38. ACOG Committee on Obstetric Practice. Gestational Hypertension and Preeclampsia: ACOG Practice Bulletin, Number 222. Obstet Gynecol. 2020;135(6):e237–e60. Epub 2020/05/23. doi: 10.1097/aog.0000000000003891. PubMed PMID: 32443079.

39. Cameron NA, Freaney PM, Wang MC, Perak AM, Dolan BM, O’Brien MJ, Tandon SD, Davis MM, Grobman WA, Allen NB, Greenland P, Lloyd-Jones DM, Khan SS. Geographic Differences in Prepregnancy Cardiometabolic Health in the United States, 2016 Through 2019. Circulation. 2022;145(7):549–51. doi: doi:10.1161/CIRCULATIONAHA.121.057107.

40. Soininen P, Kangas AJ, Würtz P, Suna T, Ala-Korpela M. Quantitative Serum Nuclear Magnetic Resonance Metabolomics in Cardiovascular Epidemiology and Genetics. Circulation: Cardiovascular Genetics. 2015;8(1):192–206. doi: doi:10.1161/CIRCGENETICS.114.000216.

41. Ussher JR, Elmariah S, Gerszten RE, Dyck JR. The Emerging Role of Metabolomics in the Diagnosis and Prognosis of Cardiovascular Disease. J Am Coll Cardiol. 2016;68(25):2850–70. Epub 2016/12/23. doi: 10.1016/j.jacc.2016.09.972. PubMed PMID: 28007146.

42. Moore BJ, White S, Washington R, Coenen N, Elixhauser A. Identifying Increased Risk of Readmission and In-hospital Mortality Using Hospital Administrative Data: The AHRQ Elixhauser Comorbidity Index. Med Care. 2017;55(7):698–705. Epub 2017/05/13. doi: 10.1097/mlr.0000000000000735. PubMed PMID: 28498196.

43. Cartus AR, Jarlenski MP, Himes KP, James AE, Naimi AI, Bodnar LM. Adverse Cardiovascular Events Following Severe Maternal Morbidity. American Journal of Epidemiology. 2022;191(1):126–36. doi: 10.1093/aje/kwab208.

44. Kleinbaum DG, Klein M. Survival analysis : a self-learning text. 3rd ed. New York: Springer; 2012. xv, 700 p. p.

45. Flanagin A, Frey T, Christiansen SL, Committee AMoS. Updated Guidance on the Reporting of Race and Ethnicity in Medical and Science Journals. JAMA. 2021;326(7):621–7. doi: 10.1001/jama.2021.13304.

46. Wenger NK, Lloyd-Jones DM, Elkind MSV, Fonarow GC, Warner JJ, Alger HM, Cheng S, Kinzy C, Hall JL, Roger VL, on behalf of the American Heart A. Call to Action for Cardiovascular Disease in Women: Epidemiology, Awareness, Access, and Delivery of Equitable Health Care: A Presidential Advisory From the American Heart Association. Circulation. 2022;145(23):e1059–e71. doi: 10.1161/CIR.0000000000001071.

47. Bandoli G, Palmsten K, Chambers CD, Jelliffe-Pawlowski LL, Baer RJ, Thompson CA. Revisiting the Table 2 fallacy: A motivating example examining preeclampsia and preterm birth. Paediatric and perinatal epidemiology. 2018;32(4):390–7. Epub 2018/05/21. doi: 10.1111/ppe.12474. PubMed PMID: 29782045.

48. Catov JM, Lewis CE, Lee M, Wellons MF, Gunderson EP. Preterm birth and future maternal blood pressure, inflammation, and intimal-medial thickness: the CARDIA study. Hypertension. 2013;61(3):641–6. Epub 2013/01/16. doi: 10.1161/hypertensionaha.111.00143. PubMed PMID: 23319540; PMCID: PMC3583341.

49. Gunderson EP, Chiang V, Pletcher MJ, Jacobs DR, Quesenberry CP, Sidney S, Lewis CE. History of gestational diabetes mellitus and future risk of atherosclerosis in mid-life: the Coronary Artery Risk Development in Young Adults study. J Am Heart Assoc. 2014;3(2):e000490. doi: 10.1161/jaha.113.000490. PubMed PMID: 24622610; PMCID: PMC4187501.

50. Gunderson EP, Sun B, Catov JM, Carnethon M, Lewis CE, Allen NB, Sidney S, Wellons M, Rana JS, Hou L, Carr JJ. Gestational Diabetes History and Glucose Tolerance After Pregnancy Associated With Coronary Artery Calcium in Women During Midlife: The CARDIA Study. Circulation. 2021;143(10):974–87. Epub 20210201. doi: 10.1161/circulationaha.120.047320. PubMed PMID: 33517667; PMCID: PMC7940578.

51. Benschop L, SchalekampDTimmermans S, Schelling SJC, Steegers EAP, Roeters van Lennep JE. Early Pregnancy Cardiovascular Health and Subclinical Atherosclerosis. Journal of the American Heart Association. 2019;8(15):e011394. doi: 10.1161/JAHA.118.011394.

52. Dodds LV, Feaster DJ, Kiefe CI, Gunderson EP, Bello NA, Rundek T, Paidas MJ, Kulandavelu S, Elfassy T. Lower Prepregnancy Cardiovascular Health is Associated With Hypertensive Disorders of Pregnancy: The CARDIA Study. Hypertension. 2025;82(10):1696–704. doi: 10.1161/HYPERTENSIONAHA.124.24517.

53. Cameron NA, Petito LC, Colangelo LA, Gunderson EP, Catov JM, Grobman WA, Rana JS, Terry JG, Lloyd-Jones DM, Allen NB, Khan SS. Prepregnancy Cardiovascular Health, Gestational Diabetes, and Coronary Artery Calcium. JAMA Cardiol. 2025;10(9):888–95. doi: 10.1001/jamacardio.2025.1887.

54. Yin J, Jin X, Shan Z, Li S, Huang H, Li P, Peng X, Peng Z, Yu K, Bao W, Yang W, Chen X, Liu L. Relationship of Sleep Duration With All-Cause Mortality and Cardiovascular Events: A Systematic Review and Dose-Response Meta-Analysis of Prospective Cohort Studies. J Am Heart Assoc. 2017;6(9). Epub 2017/09/11. doi: 10.1161/jaha.117.005947. PubMed PMID: 28889101; PMCID: PMC5634263.

55. Lu Q, Zhang X, Wang Y, Li J, Xu Y, Song X, Su S, Zhu X, Vitiello MV, Shi J. Sleep disturbances during pregnancy and adverse maternal and fetal outcomes: a systematic review and meta-analysis. Sleep medicine reviews. 2021;58:101436.

56. Hinkle SN, Schisterman EF, Liu D, Pollack AZ, Yeung EH, Mumford SL, Grantz KL, Qiao Y, Perkins NJ, Mills JL, Mendola P, Zhang C. Pregnancy Complications and Long-Term Mortality in a Diverse Cohort. Circulation. 2023;147(13):1014–25. doi: 10.1161/CIRCULATIONAHA.122.062177.

57. Aguayo L, Cotoc C, Guo JW, Labarthe DR, Allen NB, Marino BS, Davis MM, Uttal S, Lloyd-Jones DM, Perak AM. Cardiovascular Health, 2010 to 2020: A Systematic Review of a Decade of Research on Life’s Simple 7. J Am Heart Assoc. 2025;14(15):e038566. PubMed PMID: 40667836.

58. Mazzoni SE, Brewer SE, Durfee J, Pyrzanowski J, Barnard JG, Dempsey AF, O’Leary ST. Patient Perspectives of Obstetrician-Gynecologists as Primary Care Providers. The Journal of reproductive medicine. 2017;62 1–2:3–8.

59. Scholle SH, Kelleher K. Assessing Primary Care Performance in an Obstetrics/Gynecology Clinic. Women & Health. 2003;37(1):15–30. doi: 10.1300/J013v37n01_02.

60. Arendt K, Clare C, Damp J, Drummond N, D’Souza Siebert R, Ellis L, Florio K, Haddock A, Hameed A, Harris-Henderson B, Hawn J, Krening C, Lyon C, Main E, Wolfe D. Patient Safety Bundles [online resource]. 2026 [cited 2026 03/15]. Available from: https://saferbirth.org/psbs/cardiac-conditions-in-obstetric-care/.

61. Khan SS, Cameron NA, Lindley KJ. Pregnancy as an Early Cardiovascular Moment: Peripartum Cardiovascular Health. Circulation Research. 2023;132(12):1584–606. doi: 10.1161/CIRCRESAHA.123.322001.

62. Tsao CW, Aday AW, Almarzooq ZI, Anderson CAM, Arora P, Avery CL, Baker-Smith CM, Beaton AZ, Boehme AK, Buxton AE, Commodore-Mensah Y, Elkind MSV, Evenson KR, Eze-Nliam C, Fugar S, Generoso G, Heard DG, Hiremath S, Ho JE, Kalani R, Kazi DS, Ko D, Levine DA, Liu J, Ma J, Magnani JW, Michos ED, Mussolino ME, Navaneethan SD, Parikh NI, Poudel R, Rezk-Hanna M, Roth GA, Shah NS, St-Onge MP, Thacker EL, Virani SS, Voeks JH, Wang NY, Wong ND, Wong SS, Yaffe K, Martin SS, American Heart Association Council on E, Prevention Statistics C, Stroke Statistics S. Heart Disease and Stroke Statistics-2023 Update: A Report From the American Heart Association. Circulation. 2023;147(8):e93–e621. Epub 20230125. doi: 10.1161/CIR.0000000000001123. PubMed PMID: 36695182.

63. Venkatesh KK, Grobman WA, Huang X, Yee LM, Catov J, Simhan H, Haas DM, Mercer B, Reddy U, Silver RM, Levine LD, Chung J, Saade G, Greenland P, Bairey Merz CN, McNeil B, Khan SS. Association of neighborhood-level socioeconomic disadvantage and Life’s Essential 8 in early pregnancy. Am J Prev Cardiol. 2025;21:100925. Epub 20241224. doi: 10.1016/j.ajpc.2024.100925. PubMed PMID: 39838971; PMCID: PMC11750432.

64. Clark HD, Walraven Cv, Code C, Karovitch A, Keely E. Did Publication of a Clinical Practice Guideline Recommendation to Screen for Type 2 Diabetes in Women With Gestational Diabetes Change Practice? Diabetes Care. 2003;26(2):265–8. doi: 10.2337/diacare.26.2.265.

65. Ehrenthal DB, Maiden K, Rogers S, Ball A. Postpartum Healthcare After Gestational Diabetes and Hypertension. Journal of Women’s Health. 2014;23(9):760–4. doi: 10.1089/jwh.2013.4688.

66. Caraballo C, Ndumele CD, Roy B, Lu Y, Riley C, Herrin J, Krumholz HM. Trends in Racial and Ethnic Disparities in Barriers to Timely Medical Care Among Adults in the US, 1999 to 2018. JAMA Health Forum. 2022;3(10):e223856–e. doi: 10.1001/jamahealthforum.2022.3856.

67. England LJ, Grauman A, Qian C, Wilkins DG, Schisterman EF, Yu KF, Levine RJ. Misclassification of maternal smoking status and its effects on an epidemiologic study of pregnancy outcomes. Nicotine Tob Res. 2007;9(10):1005–13. Epub 2007/09/14. doi: 10.1080/14622200701491255. PubMed PMID: 17852766.

68. Stayner L, Pearce N, Nøhr E, Freeman L, Deffner V, Ferrari P, Freedman L, Kogevinas M, Kromhout H, Lewis S, MacLehose R, Parent M, Richiardi L, Shaw P, Wedekind R. Information bias: misclassification and mismeasurement of exposure and outcome. In: De González AB, Richardson DB, MK S-B, editors. Statistical Methods in Cancer Research Volume V: Bias Assessment in Case–Control and Cohort Studies for Hazard Identification. Lyon (FR): International Agency for Research on Cancer; 2024.

69. Brouwers L, van der Meiden-van Roest AJ, Savelkoul C, Vogelvang TE, Lely AT, Franx A, van Rijn BB. Recurrence of pre-eclampsia and the risk of future hypertension and cardiovascular disease: a systematic review and meta-analysis. BJOG. 2018;125(13):1642–54. PubMed PMID: 29978553.

