## Supplemental Material for "Life’s Essential 8 in Pregnancy and Time to Incident Cardiometabolic Disease Over 7 Years Follow-Up"

**Supplemental Material 1. Flow diagram of eligible Cradle Trial participants and enrollment in the PIINC cohort**

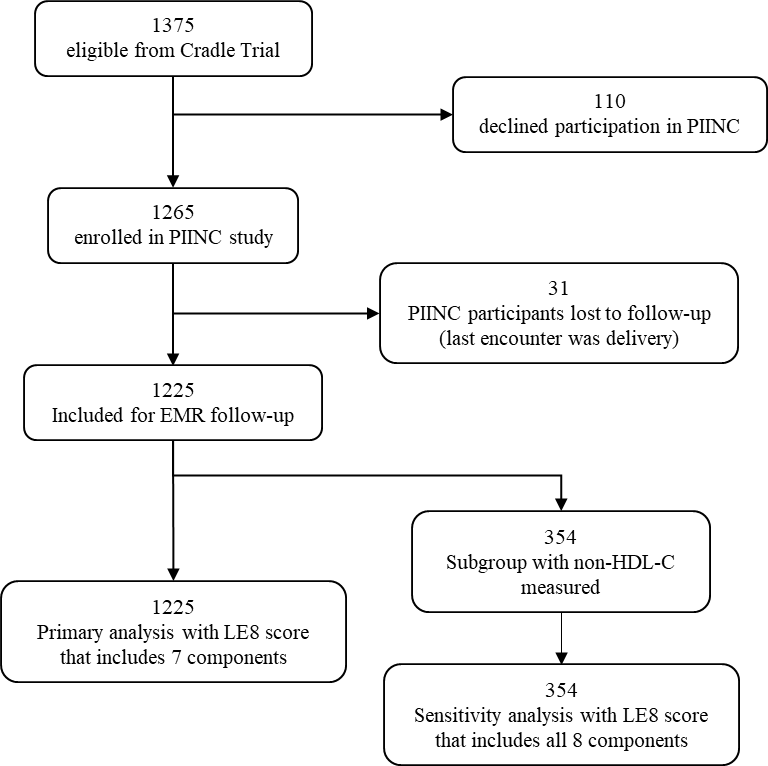

Note: All enrolled Cradle Trial participants who had not yet delivered at the time the PIINC study was initiated were eligible for participation in PIINC (N=1375)

**Supplemental Material 2: Life’s Essential Eight Scoring and Quantification of Components and Modifications.**

| **CVH metric** | **Quantification of CVH metric by AHA advisory** | | **Modifications for use in PIINC** | | |
| --- | --- | --- | --- | --- | --- |
| **Sleep** | Metric: Hours of sleep | | Survey answer to personal habits. | | |
|  | Points | Hours | Points | | “Indicate the frequency you get enough sleep” |
|  | 100 | 7–<9 | 100 | | Routinely |
|  | 90 | 9–<10 | 70 | | Often |
|  | 70 | 6–<7 |  |  |  |
|  | 40 | 5–<6 or >=10 | 40 | | Sometimes |
|  | 20 | 4–<5 | 20 | | Never |
|  | 0 | <4 |  | |  |
| **Diet** | Quantiles of HEI‐2015 (population) scoring (population) | | Modification: Food Frequency Questionnaire, adequate diet consumption – See Appendix 2. Used percentile cut-offs and scoring to align with IPAQ scoring. | | |
|  | Points | Quantile | Points | | Percentile scoring |
|  | 100 | ≥95th Percentile (top/high diet) | 100 | | ≥95^th^ P |
|  | 80 | 75th–94th Percentile | 90 | | 90-<95^th^ P |
|  |  |  | 80 | | 65-<90^th^ P |
|  | 50 | 50th–74th Percentile | 60 | | 40-<65^th^ P |
|  |  |  | 40 | | 30-<40^th^ P |
|  | 25 | 25th–49th Percentile | 20 | | 5–<30^th^ P |
|  | 0 | 1st–24th Percentile (bottom/least high quartile) | 0 | | 0-4^th^ P |
| **Physical activity** | Metric: minutes of moderate‐ to vigorous‐intensity activity per week scoring | | Modification: IPAQ. Total vigorous + moderate PA (minutes/week). | | |
|  | Points | Minutes | Points | | Percentile scoring |
|  | 100 | ≥150 | 100 | | ≥95^th^ P |
|  | 90 | 120–149 | 90 | | 90-<95^th^ P |
|  | 80 | 90–119 | 80 | | 65-<90^th^ P |
|  | 60 | 60–89 | 60 | | 40-<65^th^ P |
|  | 40 | 30–59 | 40 | | 30-<40^th^ P |
|  | 20 | 1–29 | 20 | | 5–<30^th^ P |
|  | 0 | 0 | 0 | | 0-4^th^ P |
| **Nicotine exposure** | Metric: combustible tobacco use or inhaled NDS use; | | Modification: smoking prior to and during pregnancy | | |
|  | Points | Status | Points | | Questions on smoking |
|  | 100 | Never smoker | 100 | | never smoker; |
|  | 75 | Former smoker, quit ≥5 y | 70 | | Smoking before pregnancy and No Smoking in pregnancy |
|  | 50 | Former smoker, quit 1–<5 y | 40 | | Smoking in pregnancy and No Smoking before pregnancy |
|  | 25 | Former smoker, quit <1 y, or currently using inhaled NDS | 0 | | Smoking in pregnancy and Smoking before pregnancy |
|  | 0 | Current smoker |  | |  |
| **BMI** | Metric: BMI (kg/m^2^) scoring | | No modifications. Used BMI at enrollment (12 ± 3.8 wks) | | |
|  | Points | Level |  | | |
|  | 100 | <25 |  | | |
|  | 70 | 25.0–29.9 |  | | |
|  | 30 | 30.0–34.9 |  | | |
|  | 15 | 35.0–39.9 |  | | |
|  | 0 | ≥40.0 |  | | |
| **Blood lipids** | Metric: non–HDL cholesterol (mg/dL) scoring [mmol/L to mg/dL non-HDL Cholesterol * 38.67] | | No modifications. This component was scored in the subset with metabolomics data (n=329) | | |
|  | Points | Level |  | | |
|  | 100 | <130 |  | | |
|  | 60 | 130–159 |  | | |
|  | 40 | 160–189 |  | | |
|  | 20 | 190–219 |  | | |
|  | 0 | ≥220 |  | | |
|  | If drug‐treated level, subtract 20 points | |  | | |
| **Blood glucose** | Metric: FBG (mg/dL) or HbA1c (%) scoring [28.7×HbA1c–46.7=FBG] | | Modification: Used glucose values from 1-hour 50g glucose challenge test (GCT). Back converted HbA1c for cut offs  [28.7×HbA1c–46.7=FBG]. Preserves inter-individual ranking | | |
|  | Points | Level | Points | | Level |
|  | 100 | No history of diabetes and FBG <100 (or HbA1c <5.7) | 100 | | No GDM and GCT<100 |
|  |  |  | 70 | | No GDM and GCT 100-125 |
|  | 60 | No diabetes and FBG 100–125 (or HbA1c 5.7–6.4) (prediabetes) | 60 | | No GDM and GCT 126-153 |
|  |  |  | 50 | | No GDM and GCT 154-197 (197 is maximum among non-GDM women in our study) |
|  | 40 | Diabetes with HbA1c <7.0 | 40 | | GDM with GCT <154 |
|  | 30 | Diabetes with HbA1c 7.0–7.9 | 30 | | GDM with GCT 154-180 |
|  | 20 | Diabetes with HbA1c 8.0–8.9 | 20 | | GDM with GCT 181-209 |
|  | 10 | Diabetes with Hb A1c 9.0–9.9 | 10 | | GDM with GCT 210-242 |
|  | 0 | Diabetes with HbA1c ≥10.0 | 0 | | GDM and GCT >242 |
| **Blood pressure** | Metric: systolic and diastolic blood pressures (mm Hg) Scoring | | Modification based on HDP diagnosis | | |
|  | Points | Level | Points | Level | |
|  | 100 | Systolic <120 and diastolic <80 (optimal) | 100 | No GHTN | |
|  | 75 | Systolic 120–129 and diastolic <80 (elevated) | 75 | Gestational Hypertension | |
|  | 50 | Systolic 130–139 or diastolic 80–89 (stage 1 hypertension) | 50 | Chronic Hypertension w/o PE | |
|  | 25 | Systolic 140–159 or diastolic 90–99 | 25 | PE | |
|  | 0 | Systolic: ≥160 or diastolic: ≥100 | 10 | Chronic hypertension with PE | |
|  | Subtract 20 points if treated level | |  |  | |

**Supplemental Material 3: Nutrition survey (study developed) and scoring for use in modified Life’s Essential 8**

**Prompt:** These next questions ask about certain foods you ate in the LAST MONTH.

| **Question** | **Scoring and cut offs used (ECF)** |
| --- | --- |
| About how many servings of fruit (for example apple, orange, grapes, melons, etc.), did you usually have in a day (1 serving = 1/2 cup)? | 1 cup cut-up fruit is recommended  100pts = 5 or more servings per day  50pts = 3 or 4 servings per day  25pts = 1 or 2 servings per day  0pts = zero servings (none) |
| About how many servings of vegetables (for example broccoli, spinach, greens, salad, etc.) did you usually have in a day (1 serving = 1/2 cup)? | 2.5 cups is recommended  100pts = 5 or more servings per day  50pts = 3 or 4 servings per day  25pts = 1 or 2 servings per day  0pts = zero servings (none) |
| About how many servings of grains (for example bread, cereal, rice, or pasta) did you usually have in a day (1 serving = 1 ounce)? | 3-6 ounces is recommended  100pts = 5 or more servings per day  50pts = 3 or 4 servings per day  25pts = 1 or 2 servings per day  0pts = zero servings (none) |
| About how many servings of dairy foods (for example milk, yogurt, or cheese) did you usually have in a day (1 serving = 1 cup of milk or 1 ounce of cheese)? | 3 cups milk, 1.5 ounce cheese is recommended  100pts = 3 or 4 servings per day  50pts = 5 or more servings per day  25pts = 1 or 2 servings per day  0pts = zero servings (none) |
| About how many servings of protein-containing foods (for example eggs, meat, poultry, fish, beans and peas, soy products, and nuts) did you usually have in a day (1 serving = 3 ounces of meat or 1 ounce of nuts)? | 5.5 ounces is recommended  100pts= 1 or 2 servings per day  50pts= 3 or 4 servings per day  25pts= 5 or more servings per day  0pts = zero servings (none) |
| During the last month, about how many servings of soft drinks, soda, or pop (for example, Coke, Pepsi, Sprite) did you usually have in a day (1 serving=12 ounces)? | 1 serving or less is rec  100pts= zero servings (none)  75pts= 1 or 2 servings per day  25pts= 3 or 4 servings per day  0pts= 5 or more servings per day |

Reference for cut offs of adequate consumption U.S Department of Health and Human Services and U. S Department of Agriculture, Dietary Guidelines for Americans 2015-2020. 8th ed. 2015.

Each question was scored and then the scored questions were summed. The total nutrition score for the LE8 was based on percentiles of the summed questions.

**Supplemental Material 4. ICD-10 codes for hypertensive disorders, metabolic disorders, and cardiovascular disease**

*Supplemental Material 4 is included as an excel file Supplemental Material 4.xlsx*

**Supplemental Material 5. Association of Life’s Essential 8 and Components and Time to Cardiovascular Disease Event**

| **LE8 Component score, per 10-unit increase** | **Model 1** | **P-value** | **Model 2** | **P-value** |
| --- | --- | --- | --- | --- |
|  | **Time Ratio (95%CI)** |  | **Time Ratio (95%CI)** |  |
| ***Cardiovascular disease ^c^*** | |  |  |  |
| Per 10-unit increase | 1.16 (0.87 , 1.56) | 0.32 | 1.15 (0.82 , 1.61) | 0.43 |
| High vs. low | 1.60 (0.34, 7.52) | 0.55 | 1.38 (0.28, 6.86) | 0.69 |
| Moderate vs. low | 1.87 (0.46, 7.50) | 0.38 | 2.31 (0.56, 9.51) | 0.25 |
| ***Cardiovascular disease ^c^*** |  |  |  |  |
| Glucose | 0.59 (0.32, 1.08) | 0.09 | 0.53 (0.24, 1.21) | 0.13 |
| BMI | 0.99 (0.71, 1.39) | 0.96 | 0.90 (0.57, 1.43) | 0.65 |
| Smoking | 1.24 (0.93, 1.66) | 0.15 | 1.27 (0.84, 1.91) | 0.26 |
| Hypertension | 1.69 (1.11, 2.58) | 0.01 | 2.03 (1.14, 3.60) | 0.02 |
| Physical activity | 1.14 (0.47, 2.77) | 0.77 | 0.90 (0.11, 7.33) | 0.92 |
| Diet | 1.28 (0.74, 2.22) | 0.38 | 0.94 (0.35, 2.53) | 0.90 |
| Sleep | 1.50 (0.90, 2.50) | 0.12 | 1.89 (0.90, 3.99) | 0.09 |

Model 1: adjusted for maternal age

Model 2: adjusted for maternal age, ethnicity (Hispanic vs. non-Hispanic), gravidity (continuously), education, income, insurance.
